# Beyond Agrammatism: Syntax Processing in Chinese Primary Progressive Aphasia

**DOI:** 10.64898/2026.10.16.26365322

**Authors:** Yu-Wen Cheng, Ta-Fu Chen, Lorinda Kwan-Chen Li-Ying, Raymond Y. Lo, Joshua Tsoh, Andrew Lung-Tat Chan, Adrian Wong, Vincent Chung Tong Mok, Connie TY Yan, Chien Jung Lu, Yu Sun, Pei-Ning Wang, YiChen Lee, Adeline Ng, FangDa Leng, Yu Chen, Cheng-Hsuan Li, Chin-Hsien Lin, Yu-Chen Chuang, Maria-Luisa Mandelli, Maria Luisa Gorno-Tempini, Boon Lead Tee

## Abstract

Syntax processing impairment is a defining feature of primary progressive aphasia (PPA), particularly in the nonfluent/agrammatic variant. However, current models are derived largely from Indo-European languages, and it remains unclear whether the cognitive and neuroanatomical mechanisms of syntax processing generalize to Chinese, an analytic, pro-drop language that encodes grammatical relationships primarily through word order and function words rather than inflectional morphology.

Participants were recruited from the international Chinese Language Assessment in PPA cohort and included 64 individuals with PPA (19 logopenic, 15 nonfluent/agrammatic, 15 semantic, and 15 unclassifiable/mixed) and 60 cognitively healthy controls. Participants completed sentence comprehension and production tasks with systematically manipulated syntactic features. Item-level linguistic determinants were examined using ridge regression, diagnostic performance using receiver operating characteristic analyses, and neuroanatomical correlates using voxel-based morphometry of structural MRI.

Performance on sentence comprehension and production was influenced by grammatical and lexical features. Subject-relative clauses, passive constructions, a higher proportion of function words, longer character length, and greater lexical content were associated with poorer performance among participants with PPA. Sentence comprehension was impaired in the logopenic, nonfluent/agrammatic, and semantic variants, even when isolated to the syntactic component of the sentence comprehension task. Similarly, sentence production was impaired across all PPA variants (all *P* < 0.01 versus controls). No significant differences were observed among PPA variants. Adjusted receiver operating characteristic analyses demonstrated high discrimination between each PPA variant and controls, with areas under the curve ranging from 0.87 to 0.97 for sentence comprehension-syntax and from 0.91 to 0.97 for sentence production. However, pairwise comparisons showed no significant differences in discriminative performance among PPA variants after correction for multiple comparisons. Both sentence comprehension-syntax and sentence production showed associations with semantic and repetition measures in addition to motor speech measures. Voxel-based morphometry showed that higher sentence comprehension-syntax scores were associated with greater grey matter volume in left frontotemporal and cingulate regions, whereas higher sentence production scores were additionally associated with greater gray matter volume in frontostriatal structures, including the caudate, putamen, and thalamus.

Syntactic processing impairment in Chinese-speaking individuals with PPA extends beyond the nonfluent/agrammatic variant and is associated with distributed frontotemporal and frontostriatal regions, as well as lexical-semantic and auditory verbal short term memory mechanisms. These findings suggest that the cognitive and neural manifestations of syntactic impairment may be shaped by language-specific typological properties, underscoring the need for linguistically informed approaches to the assessment and interpretation of PPA.

## Introduction

Primary progressive aphasia (PPA) is a neurodegenerative syndrome characterized by progressive deterioration of speech and language functions, and is typically classified into three canonical variants, the nonfluent/agrammatic (nfvPPA), logopenic (lvPPA), and semantic (svPPA) variants.^1^ The nonfluent/agrammatic variant is characterized by effortful, halting speech and/or agrammatism, typically associated with left frontal-insular degeneration. The semantic variant is characterized by impaired confrontation naming and single-word comprehension, reflecting loss of semantic knowledge and left anterior temporal lobe atrophy. The logopenic variant is characterized by word-finding difficulties, impaired sentence repetition, and phonological errors with relatively preserved single-word comprehension, typically associated with degeneration of the left temporoparietal junction.^1,2^

Owing to its defining agrammatic features, nfvPPA is classically characterized by impaired processing of syntactically complex sentences. Consequently, individuals with nfvPPA often demonstrate deficits in syntax processing across both language comprehension and production. Sentence comprehension is disproportionately impaired for syntactically complex, non-canonical sentence structures, while speech production is characterised by agrammatism, including shortened phrase length, omission of grammatical morphemes, and reduced use of complex syntactic constructions^3^. Prior work using the Northwestern Assessment of Verbs and Sentences (NAVS) has demonstrated that deficits in syntactic comprehension and production are highly correlated in individuals with nfvPPA, suggesting disruption of shared core grammatical processes.^4,5^ Structural and functional neuroimaging studies have further implicated the left inferior frontal cortex as a key component of the distributed neural network supporting syntactic comprehension and production.^6,7^ Voxel-based morphometry studies consistently associate atrophy of this region with receptive and expressive syntax processing deficits in nfvPPA, while functional MRI reveals that left inferior frontal cortex fails to modulate activity in response to syntactic complexity, a response observed in healthy controls and largely preserved in svPPA, where syntax processing is relatively spared.^8–10^

Current understanding of PPA has been derived predominantly from studies of English-speaking individuals and speakers of other Indo-European languages.^11,12^ This reliance has constrained both the theoretical scope and clinical applicability of existing PPA models, as language typology is increasingly recognised to shape the manifestation of language impairment in PPA.^13,14^ Cross-linguistic comparisons between English and Italian, for example, have shown that the clinical manifestations of PPA may differ across languages. Among patients with nfvPPA, expressive agrammatism is more prevalent in Italian, a synthetic language with rich inflectional morphology and flexible word order, whereas English-speaking patients more frequently exhibit speech sound distortions, potentially reflecting the greater articulatory demands of English.^13,14^ These findings raise the possibility that PPA variants that affect similar speech and language networks do not necessarily produce a uniform clinical phenotype across languages; rather, the structural properties of individual languages may influence which aspects of grammar are most susceptible to disruption. Cross-linguistic investigation is therefore essential for distinguishing universal from language-specific mechanisms of syntactic impairment and for ensuring that diagnostic criteria and assessment tools remain valid across typologically diverse languages and populations.^8^

Chinese provides a particularly informative test case because it differs fundamentally from the Indo-European languages on which current models are based. Whereas English encodes grammatical information partly through inflectional morphology, including tense and agreement marking, Chinese is a predominantly analytic language with minimal inflectional morphology, relying instead on word order, function words, aspectual particles, and contextual information to signal grammatical relationships.^15^ Consequently, syntactic structure in Chinese is realized primarily through word order and function words rather than morphological inflection. These typological characteristics raise the possibility that processing syntactically complex sentences in Chinese may depend not only on grammatical computation but also on lexicosemantic integration and auditory verbal short term memory, particularly when syntactic complexity is accompanied by increased sentence length and a greater number of lexical-semantic units. Accordingly, in Chinese-speaking populations, deficits in syntax comprehension and production may not be restricted to nfvPPA but may also manifest in lvPPA and svPPA through disruption of the cognitive processes required to maintain and integrate linguistic information. Furthermore, the neural substrates associated with syntactic performance in Chinese may extend beyond the regions most consistently implicated in studies of predominantly English-speaking populations. Whether the cognitive and neuroanatomical mechanisms underlying impairments in syntax processing in PPA generalize across these typologically distinct languages therefore remains an important unresolved question.

To address these questions, we investigated syntax processing in the international Chinese Language Assessment for Primary Progressive Aphasia (CLAP) cohort of Chinese-speaking individuals with PPA and cognitively healthy controls. Using sentence comprehension and production tasks with systematically manipulated syntactic complexity, we examined: (i) the linguistic factors associated with syntax task performance; (ii) differences in syntax processing performance across PPA variants and the diagnostic utility of these tasks for distinguishing PPA from cognitively healthy controls; and (iii) the neuroanatomical correlates of impairments in syntax processing using voxel-based morphometry. Accordingly, we hypothesized that the analytic structure of Chinese would shape the cognitive and neural determinants of syntax processing. Specifically, we predicted that syntax processing performance would depend not only on grammatical complexity but also on lexical-semantic and auditory verbal short term memory demands, resulting in impairments across multiple PPA variants and engaging neural regions beyond those previously reported in predominantly English-speaking patients with PPA.

## Materials and methods

### Study participants and language assessment

A total of 124 Mandarin- and Cantonese-speaking participants with PPA and cognitively healthy controls were recruited through the international Chinese Language Assessment in PPA (CLAP) project.^16^ The CLAP project is a prospective, multicenter cohort study involving seven sites across Hong Kong, Taiwan, and the United States. Participants were required to have completed at least six years of formal education in Mandarin or Cantonese languages; those with fewer than six years of schooling were eligible if all formal education was conducted in Mandarin or Cantonese languages. Exclusion criteria included other major neurodegenerative disorders, prior brain surgery, major traumatic brain injury requiring hospitalization, oropharyngeal conditions affecting articulation, severe visual or hearing impairment, or a Mini–Mental State Examination (MMSE)^17^ score lower than 10. All participants underwent standardized clinical, neuropsychological, and neuroimaging assessments according to the CLAP study protocol, including the MMSE, the CLAP language battery, and brain MRI. Individuals who completed the sentence comprehension and/or production tasks were included in the present study. Neuropsycholinguistic assessments were administered by board-certified neurologists, neuropsychologists, speech–language pathologists, or supervised research staff, all of whom were native speakers of Mandarin or Cantonese and proficient in both language varieties. As previously described,^16,18^ the CLAP language battery comprises a comprehensive assessment of speech and language, including measures of confrontation naming, single-word comprehension, semantic association, repetition, and motor speech. All assessments were conducted in each participant’s dominant language (Mandarin or Cantonese), as determined by a language proficiency and use questionnaire.^19^

### Standard protocol approvals, registrations, and patient consents

The study protocol was approved by the institutional review boards or research ethics committees of all participating institutions. Written informed consent was obtained from all participants or their legally authorized representatives prior to study enrollment.

### Sentence comprehension and production tasks

The sentence comprehension and production tests were developed based on the testing framework of Northwestern Assessment of Verbs and Sentences (NAVS) test.^20^ The task structure is illustrated in Fig. 1A. In the sentence comprehension (SC) task, participants were presented with 30 Chinese sentences describing interactions between two characters, with syntactic complexity systematically varied across items. Each sentence was accompanied by two or four corresponding illustrations, and participants were tasked to select the picture that best matched the sentence meaning. For syntactic-foil items, the picture alternatives differed in the assignment of thematic roles among the depicted characters, such that correct responses depended on interpreting word order and function words. In contrast, for semantic-foil items, the alternatives differed in the lexicosemantic information conveyed by the sentence. Examples of syntactic- and semantic-foil items are shown in Supplementary Fig. 1. The total SC score comprised all 30 items, including 23 syntactic-foil items (SC-syntax) and 7 semantic-foil items (SC-semantics).

**Figure 1.**
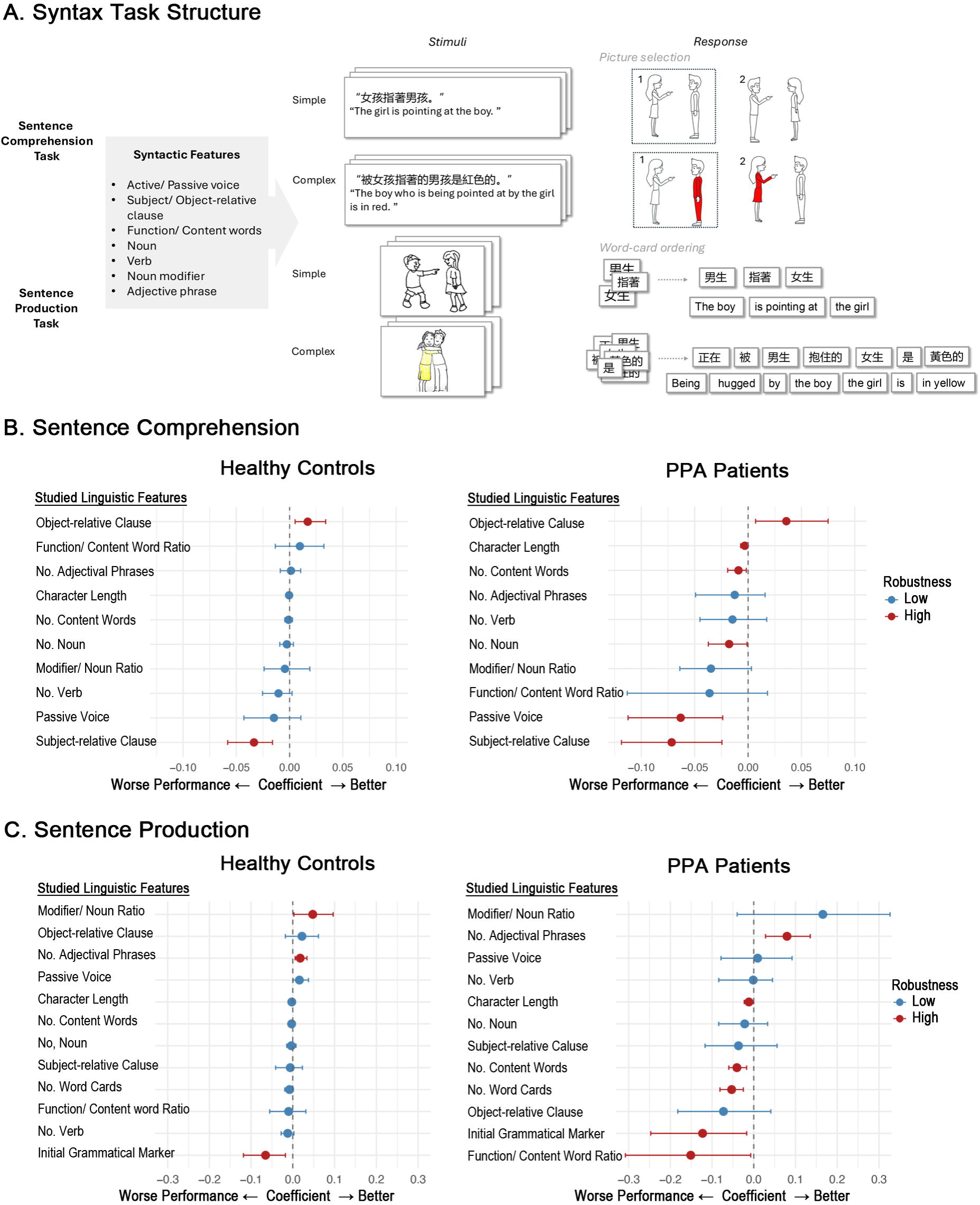
Task structure and syntactic predictors of test performance in healthy control and patients with primary progressive aphasia. (**A**) Structure of the sentence comprehension and sentence production tasks. **(B)** Syntactic features associated with sentence comprehension performance in healthy controls and patients with primary progressive aphasia (PPA). **(C)** Syntactic features associated with sentence production performance in the participant groups. Across panels B and C, feature importance was estimated using ridge regression models. High-robustness features denote predictors that achieved statistical significance under both bootstrap resampling and principal component (PC)-based inference, whereas low-robustness features indicate predictors not consistently supported by both methods. Abbreviation: PPA = Primary Progressive Aphasia.

In the sentence production (SP) task, participants were presented with pictures depicting interactions between two characters and instructed to construct a grammatically well-formed sentence using a set of three to seven word cards provided for each stimulus. To minimize response ambiguity, the first word of the target sentence was provided. A total of 36 items were administered for the SP task. Across both SC and SP tasks, each stimulus was annotated for a predefined set of linguistic features spanning three domains: syntactic structure, lexical composition, and task design features (Figure 1A and Table 1). Syntactic features included sentence voice (active vs. passive) and relative clause type (subject-vs. object-relative). For example, the subject-relative construction ‘正在推女孩的男孩是紅色的’ (‘The boy who is pushing the girl is red’) contrasts with the object-relative construction ‘女孩正在推的男孩是 紅色的’ (‘The boy whom the girl is pushing is red’), in which the modified noun functions as the subject and object of the relative clause, respectively. Lexical features included sentence length (character count), number of content words, number of function words, function-to-content word ratio, number of nouns and verbs, noun modifiers, and adjectival phrases. For the task design features, two additional task-specific features were evaluated: the number of word cards and the lexical category of the initial word card, which could be either a grammatical marker (e.g., the progressive aspect marker 正在 [is/are] or the passive marker 被 [be]) or a lexical content word (e.g., 男孩, ‘boy’). Accuracy was scored as the proportion of correct responses (0-30/30 for SC; 0-36/36 for SP), with higher scores indicating better performance.

### Statistical analysis

Clinical and demographic characteristics across diagnostic groups were compared using one-way analysis of variance (ANOVA) for continuous variables and chi-square (χ²) tests for categorical variables, with Tukey’s post hoc test for pairwise comparisons. Because SC and SP scores were right-skewed and exhibited ceiling effects particularly among cognitively healthy controls, group differences in task performance were evaluated using the nonparametric Kruskal-Wallis test followed by Bonferroni-corrected pairwise comparisons.

To identify syntactic features associated with item-level accuracy, ridge regression (L2-regularized linear regression) models were fitted using the *glmnet* and *ridge*^21^ packages in R. Item-level analyses included all 30 SC items to capture the full range of semantic and syntactic manipulations. The regularization parameter (λ) was selected using 10-fold cross-validation. Coefficient stability was evaluated using 1,000 bootstrap resamples to derive 95% confidence intervals, with predictors whose intervals excluded zero considered reliable. Principal component–based *p* values from *ridge::linearRidge* were additionally reported based on the inference framework described by Cule *et al.*,^22^ and predictors meeting significance criteria under both approaches were classified as high-robustness features.

Given the limited variability in the 7-item SC-semantics subscore and the primary focus of this study on syntax processing, subsequent analyses focused on SC-syntax, defined as the 23-item syntactic-foil subscore of the 30-item SC task (range, 0-23), and SP, defined as the total score of the 36-item SP task (range, 0-36). The diagnostic performance of SC-syntax and SP in discriminating each PPA variant from cognitively healthy controls was assessed using receiver operating characteristic (ROC) curve analyses. For each analysis, participants with a given PPA variant were compared with controls. ROC analyses were conducted using both univariable models based on the SC-syntax and SP scores alone and multivariable logistic regression models adjusted for age, sex, and years of education.

We next examined whether the ability to discriminate each PPA variant from cognitively healthy controls differed across variants. To further assess this, AUCs from the covariate-adjusted variant-versus-control ROC models were compared pairwise using a generalized DeLong method accounting for covariance arising from the shared cognitively healthy control group. Six prespecified pairwise comparisons among the four variant-specific AUCs were performed separately for SC-syntax and SP, with Bonferroni correction for multiple comparisons.

Finally, to further evaluate the variant specificity of syntax processing performance, additional univariable pairwise ROC analyses were performed across all diagnostic groups using SC-syntax and SP scores alone. Pairwise discriminative performance was summarized as heatmaps displaying the area under the ROC curve (AUC) for each diagnostic comparison. All statistical tests were two-sided, and a *p* value < 0.05 was considered statistically significant. Analyses were performed using R version 4.2.3.

### Neuroimaging acquisition and analysis

To examine the neuroanatomical correlates of SC-syntax and SP, voxel-based morphometry (VBM) analyses were conducted on structural MRI scans acquired within three months of the syntax tasks and other CLAP battery assessments. Structural brain MRI scans were acquired on 3 Tesla scanners using the standardized Alzheimer’s Disease Neuroimaging Initiative 3 (ADNI-3; http://adni.loni.usc.edu/adni-3/) imaging protocol. High-resolution T1-weighted anatomical images were obtained with a three-dimensional magnetization-prepared rapid acquisition gradient-echo (MPRAGE) sequence. Acquisition parameters included 200–211 sagittal slices, repetition time = 2300 ms, echo time = 2.98 ms, inversion time = 900 ms, flip angle = 9° to 11°, field of view = 256 × 256 mm, matrix size = 256 × 256, in-plane voxel size of 1.0 × 1.0 mm^2^, and slice thickness = 1 mm.

Imaging preprocessing and VBM analyses were conducted using the Computational Anatomy Toolbox (CAT12) implemented in Statistical Parametric Mapping (SPM12; Wellcome Trust Centre for Neuroimaging), running under MATLAB R2022b. Structural T1-weighted images were first visually inspected for motion and other artifacts, followed by bias-field correction, skull stripping, and tissue segmentation into gray matter, white matter, and cerebrospinal fluid using CAT12’s default tissue probability maps. Gray matter probability maps were then spatially normalized to Montreal Neurological Institute (MNI) space using CAT12’s high-dimensional diffeomorphic registration (geodesic shooting) algorithm, modulated by the Jacobian determinants to preserve regional volume information, and smoothed with an isotropic Gaussian kernel of 8-mm full-width at half-maximum (FWHM).

Voxel-wise multiple regression analyses were performed separately to examine associations between gray matter volume and SC-syntax and SP scores, representing syntax processing of sentence comprehension and production, respectively. Age, sex, and total intracranial volume (TIV) were included as covariates in all models. Statistical significance for the primary VBM analyses was thresholded at a voxel-wise family-wise error (FWE)-corrected significance level of *P* < 0.05. Only clusters with an extent greater than 100 voxels (*k* > 100) were reported. Because of the limited sample size in the PPA variant-specific analyses, these exploratory results are presented at an uncorrected voxel-wise threshold of *P* < 0.0001 with a minimum cluster extent of *k* > 200 voxels for visualization purposes.

## Results

### Clinical profiles of participants

Sixty-four individuals with PPA, including 19 with lvPPA, 15 with nfvPPA, 15 with svPPA, and 15 with unclassifiable or mixed PPA, along with 60 cognitively healthy control participants were enrolled for this study. Among these participants, 114 (56 individuals with PPA and 58 controls) completed the sentence comprehension test, whereas 81 (43 individuals with PPA and 38 controls) completed the sentence production test. Demographic and neuropsychological characteristics are summarized in Table 2. Compared with cognitively healthy controls, individuals with lvPPA were older (69.8 ± 7.3 vs. 63.5 ± 8.4 years; post-hoc adjusted *P* = 0.049) and were less frequently male (32% vs. 68%; post-hoc adjusted *P* = 0.025). Educational attainment was significantly lower in lvPPA (11.6 ± 4.1 years), nfvPPA (12.4 ± 4.1 years), svPPA (13.8 ± 3.1 years), and unclassifiable or mixed PPA (12.7 ± 4.5 years), compared with controls (15.9 ± 3.3 years; all post-hoc adjusted *P* < 0.05). Global cognitive performance, as measured by the MMSE, was reduced across all PPA variants (lvPPA: 19.8 ± 4.0; nfvPPA: 24.9 ± 4.8; svPPA: 21.2 ± 4.2; unclassifiable/mixed PPA: 24.3 ± 4.3), relative to controls (29.2 ± 1.2; all post-hoc adjusted *P* < 0.01). There were no significant group differences in handedness or the distribution of test languages.

Across speech and language assessments, all PPA variants demonstrated impaired performance relative to cognitively healthy controls, with distinct subtype-specific profiles. Participants with lvPPA showed reduced repetition accuracy compared with controls (59.8 ± 20.9% vs. 89.6 ± 5.4%, post-hoc adjusted *p* < 0.0001) and those with unclassifiable or mixed PPA (post-hoc adjusted *P* = 0.049). Participants with svPPA exhibited marked impairments in single word comprehension and semantic association tasks relative to controls (single-word comprehension: 7.0 ± 3.3 vs. 14.6 ± 0.6, *P* < 0.0001; semantic association picture: 9.4 ± 3.6 vs. 14.9 ± 0.3, *p* < 0.0001; semantic association word: 9.2 ± 3.4 vs. 14.8 ± 0.4, *P* < 0.0001) and all other PPA variants (all post-hoc adjusted *P* < 0.0001). In contrast, participants with nfvPPA showed prominent deficits on both motor speech measures compared with controls (manner of articulation: 53.2 ± 27.1 vs. 92.2 ± 8.9, *p* < 0.0001; tone of articulation: 56.0 ± 30.0 vs. 93.0 ± 8.1, *p* < 0.0001) and the other PPA groups (all post-hoc adjusted *P* < 0.001).

### Syntactic features associated with sentence comprehension and production performance

To identify linguistic features associated with SC and SP performance, ridge regression analyses were performed separately in participants with PPA and cognitively healthy controls, allowing comparison of the determinants of performance under healthy and disease conditions. The results of these analyses are presented in Fig. 1B and C for SC and SP, respectively. Syntactic and lexical features that reached statistical significance under both bootstrap resampling and principal component (PC) based inference were classified as “high-robustness” predictors.

For the SC task, sentences containing subject-relative clauses were associated with lower accuracy (i.e., worse performance), whereas sentences containing object-relative clauses were associated with higher accuracy (i.e., better performance) among cognitively healthy controls (Fig. 1B). A similar pattern was observed in participants with PPA, but with larger effect sizes. In addition, among participants with PPA, sentences featuring passive voice in the main clause, a greater number of nouns, a higher number of content words, and longer character length were further associated with reduced comprehension accuracy.

For the SP task, sentences in which the initial word card was a grammatical marker rather than content word were associated with lower accuracy in cognitively healthy controls, whereas sentences containing a greater number of adjective phrases or a higher modifier-to-noun ratio were associated with better performance (Fig. 1C). The associations between SP and both initial word type and the number of adjectival phrases were preserved in participants with PPA. In addition, poorer production performance was associated with a higher function-to-content word ratio, a greater number of word cards, more content words, and longer character length.

### Performance on sentence comprehension and production across PPA variants

Performance on the SC and SP tasks across diagnostic groups is presented in Fig. 2. For the SC task, individuals with lvPPA, nfvPPA, and svPPA demonstrated significantly lower performance than cognitively healthy controls (lvPPA: 24.4 ± 3.6; nfvPPA: 25.4 ± 4.9; svPPA: 26.1 ± 2.7; controls: 29.2 ± 1.2; all *P* < 0.01 vs. controls; Fig. 2A). The unclassifiable/mixed PPA group did not differ significantly from controls. Analysis of the two SC subscores showed a similar pattern for SC-syntax, with individuals with lvPPA, nfvPPA, and svPPA scoring significantly lower than controls (lvPPA: 17.9 ± 3.2; nfvPPA: 19.3 ± 3.6; svPPA: 20.3 ± 2.1; controls: 22.4 ± 1.1; all *P* < 0.01 vs. controls; Fig. 2C). In contrast, for SC-semantics, only individuals with svPPA performed significantly worse than cognitively healthy controls (lvPPA: 6.4 ± 0.8; nfvPPA: 6.1 ± 1.6; svPPA: 5.9 ± 0.9; controls: 6.8 ± 0.5; *P* < 0.0001 for svPPA vs. controls; Fig. 2D). For the SP task, individuals across all PPA variants demonstrated lower performance than cognitively healthy controls (lvPPA: 23.4 ± 9.7; nfvPPA: 28.4 ± 5.4; svPPA: 25.6 ± 7.5; unclassifiable/mixed PPA: 24.5 ± 7.9; controls: 35.0 ± 1.6; all *P* < 0.01 vs. controls; Fig. 2B). No statistically significant differences were observed among different PPA variants for either SC or SP performance.

**Figure 2.**
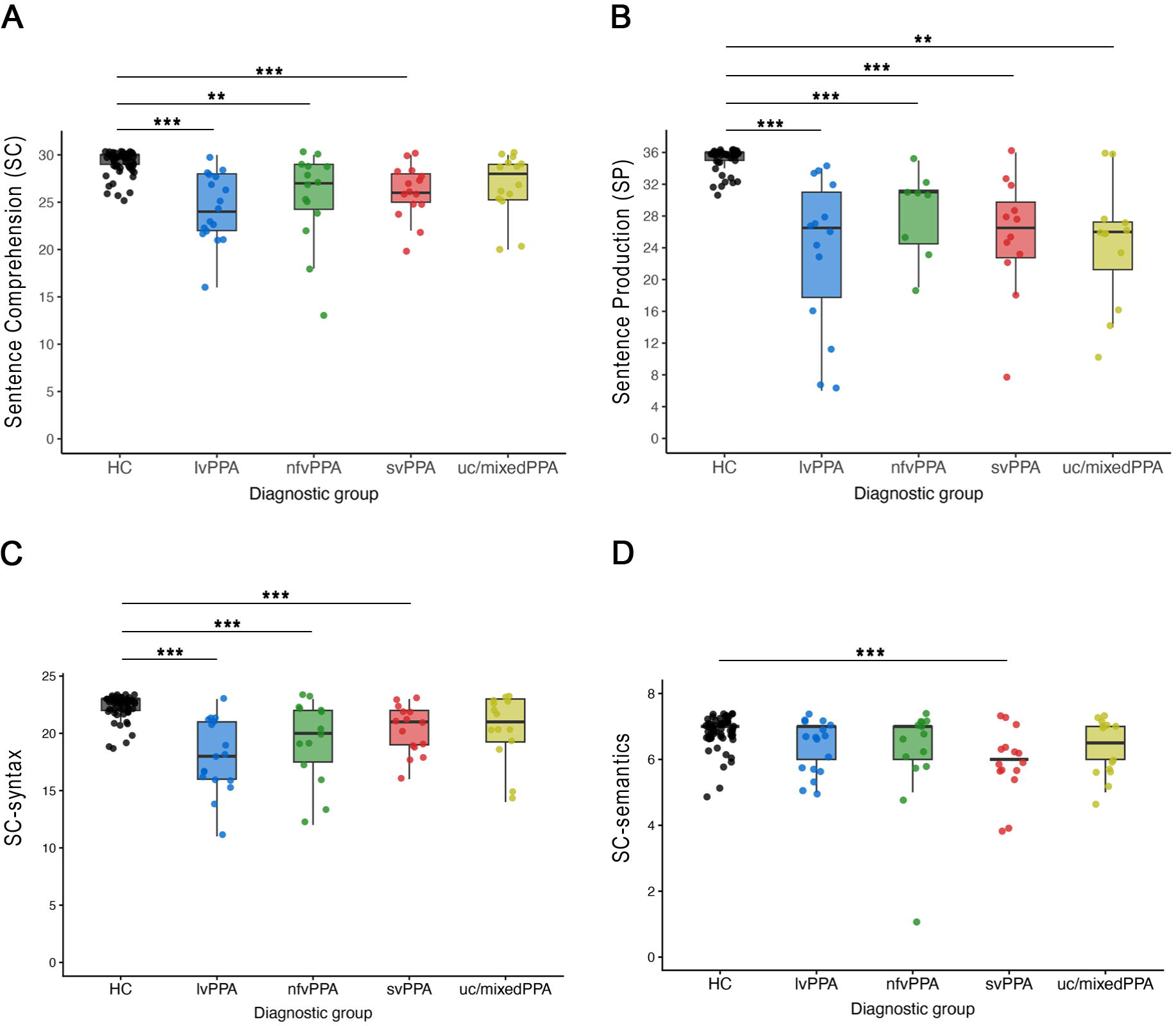
Sentence Comprehension and Production Performance Across PPA Variants. **(A)** Sentence comprehension (SC), based on the total score of the 30-item sentence comprehension task. **(B)** Syntax production (SP), based on the total score of the 36-item syntax production task. **(C)** SC-syntax, based on the 23-item syntactic-foil subscore of the SC task. **(D)** SC-semantics, based on the 7-item semantic-foil subscore of the SC task.. Boxplots represent the median, interquartile range, and range of scores; individual data points are shown. Asterisks indicate significant differences compared with healthy controls (** = p < 0.01; *** = p < 0.001). In panel D, the box is not visible for some groups because the first and third quartiles are identical (IQR = 0), indicating that the middle 50% of scores are concentrated at the same value. Note: the SC analyses included 114 participants (17 lvPPA, 14 nfvPPA, 15 svPPA, 10 unclassifiable/mixed PPA, and 58 HC), whereas the SP analyses included 81 participants (14 lvPPA, 8 nfvPPA, 12 svPPA, 9 unclassifiable/mixed PPA, and 38 HC). Abbreviations: HC = healthy control; lvPPA = logopenic variant primary progressive aphasia; nfvPPA = nonfluent/agrammatic variant primary progressive aphasia; PPA = primary progressive aphasia; ROC = receiver operating characteristic; svPPA = semantic variant primary progressive aphasia; uc/mixedPPA = unclassifiable/mixed primary progressive aphasia.

### Diagnostic discrimination of PPA variants by syntax measures

ROC curve analyses were performed to evaluate the ability of SC-syntax and SP scores to discriminate each PPA variant from cognitively healthy controls. As age, sex, and educational attainment differed across diagnostic groups, primary analyses used covariate-adjusted ROC models accounting for these variables (Fig. 3A and 3B). Univariable ROC analyses based on SC-syntax and SP scores alone are provided in Supplementary Fig. 2. Covariate-adjusted ROC analyses demonstrated high discrimination of each PPA variant from cognitively healthy controls for both the SC-syntax and SP (Fig. 3A and 3B). For SC-syntax, discriminative performance was highest for lvPPA (AUC = 0.97, 95% CI: 0.94, 1.00), followed by nfvPPA (AUC = 0.89, 95% CI: 0.81, 0.98), svPPA (AUC = 0.87, 95% CI: 0.76, 0.98), and unclassifiable/mixed PPA (AUC = 0.87, 95% CI: 0.78, 0.96). For SP, discriminative performance was highest for lvPPA and unclassifiable/mixed PPA (both AUC = 0.97; lvPPA: 95% CI: 0.94, 1.00; unclassifiable/mixed PPA: 95% CI: 0.92, 1.00), followed by nfvPPA (AUC = 0.95, 95% CI: 0.89, 1.00) and svPPA (AUC = 0.91, 95% CI: 0.78, 1.00). Univariable ROC analyses yielded a similar pattern of discriminative performance across PPA variants (Supplementary Fig. 2), with AUCs ranging from 0.70 to 0.93 for SC-syntax and from 0.89 to 0.95 for SP. Pairwise comparisons of the covariate-adjusted ROC curves using a generalized DeLong method showed no statistically significant differences in discriminative performance among PPA variants for either SC-syntax or SP after Bonferroni correction for six pairwise comparisons (adjusted *α* = 0.0083).

**Figure 3.**
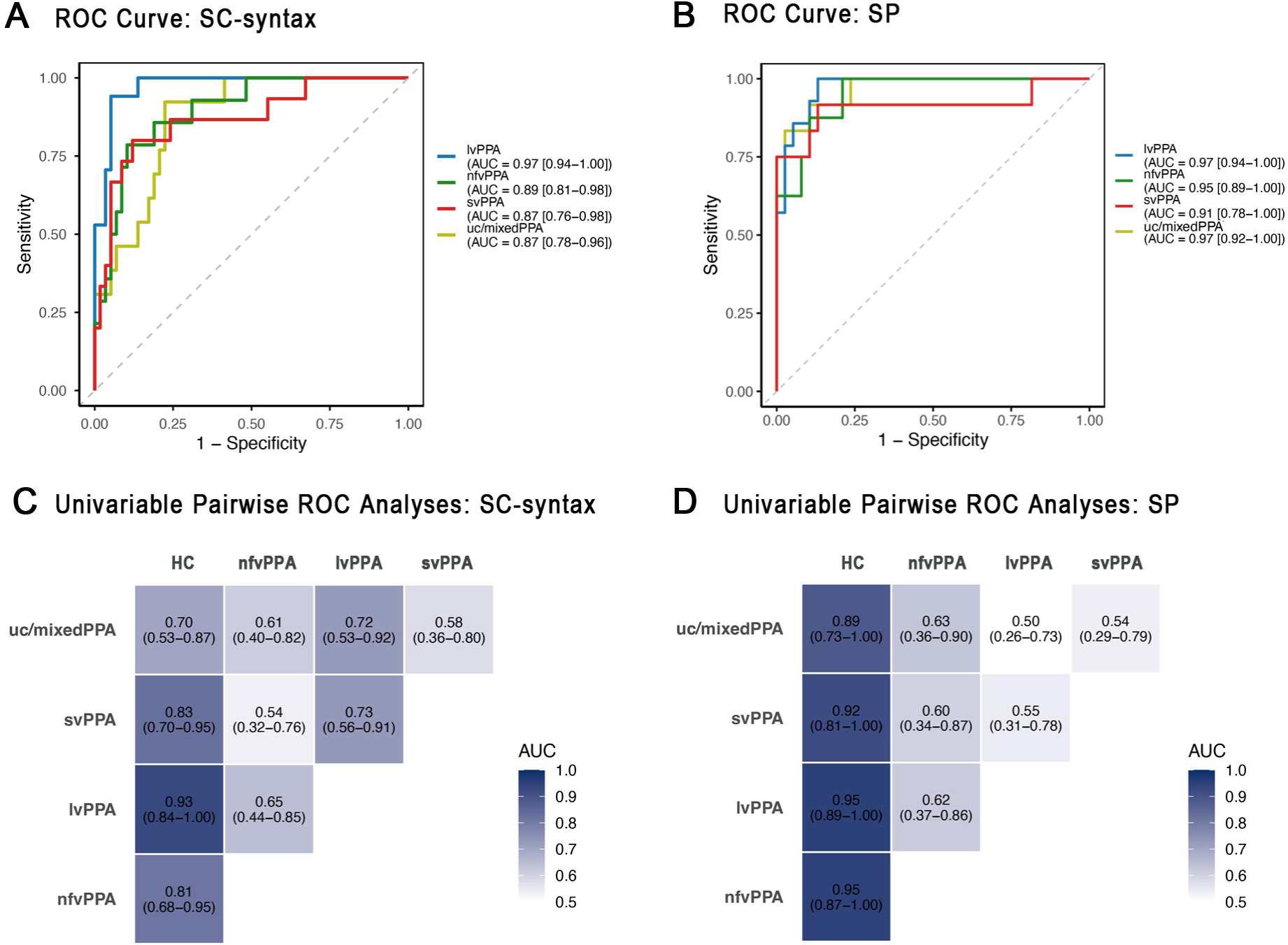
Diagnostic Performance of SC-Syntax and SP Measures Among PPA Variants. **(A-B)** Receiver operating characteristic (ROC) curves illustrating the performance of the SC-syntax **(A)** and SP **(B)** scores in distinguishing each PPA subtype from healthy controls. ROC analyses were adjusted for age, sex, and years of education. **(C-D)** Heatmaps summarizing pairwise univariable ROC analyses based on SC-syntax **(C)** and SP **(D)** scores for all diagnostic group comparisons. Each cell displays the area under the ROC curve (AUC) with the corresponding 95% confidence interval, illustrating the discriminative performance of syntax tasks between pairs of diagnostic groups. Abbreviations: AUC = area under the curve; CI = confidence interval; HC = healthy control; lvPPA = logopenic variant primary progressive aphasia; nfvPPA = nonfluent/agrammatic variant primary progressive aphasia; PPA = primary progressive aphasia; ROC = receiver operating characteristic; svPPA = semantic variant primary progressive aphasia; uc/mixedPPA = unclassifiable/mixed primary progressive aphasia.

To examine whether syntax performance showed specificity for nfvPPA, pairwise ROC analyses based on SC-syntax and SP scores alone were performed between all diagnostic groups (Fig. 3C and 3D). Although both measures showed good discrimination between each PPA variant and healthy controls, their ability to distinguish among PPA variants was substantially lower. For SC-syntax, pairwise discrimination between nfvPPA and lvPPA, svPPA, and unclassifiable/mixed PPA was limited (AUCs = 0.54-0.65). Similarly, SP showed limited discrimination between nfvPPA and the other PPA variants (AUCs = 0.60-0.63).

To further characterize the cognitive processes underlying syntax task performance, we examined correlations between SC-syntax and SP scores and measures of single-word comprehension, semantic association, repetition, and motor speech. Both SC-syntax and SP scores showed moderate-to-strong positive correlations with single-word comprehension, semantic association, and repetition (Spearman’s *ρ* = 0.49-0.61; Supplementary Fig. 3). Correlations with motor speech measures were moderate (Spearman’s ρ = 0.39-0.54).

### Neuroanatomical correlations of syntax processing

VBM analyses revealed significant associations between regional grey matter volume and SC-syntax and SP performance (Fig. 4; Supplementary Tables 1 and 2). The SC-syntax analysis included 79 participants (14 lvPPA, 8 nfvPPA, 9 svPPA, 7 unclassifiable/mixed PPA, and 41 cognitively healthy controls), while the SP analysis included 60 participants (10 lvPPA, 6 nfvPPA, 7 svPPA, 6 unclassifiable/mixed PPA, and 31 cognitively healthy controls).

**Figure 4.**
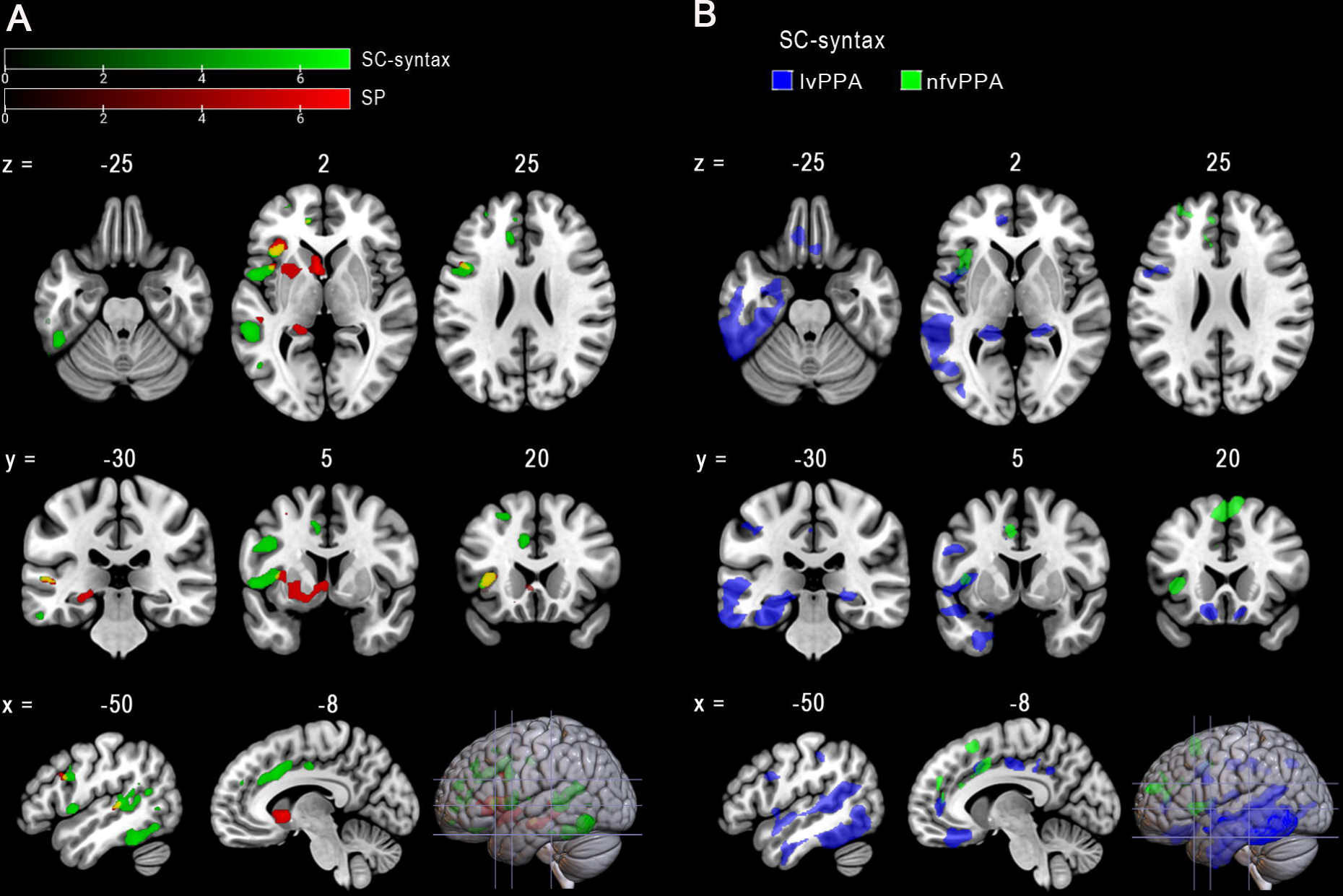
Neuroanatomical Correlations of SC-Syntax and SP Measures. **(A)** Voxel-based morphometry analyses showing regions in which lower gray matter volume was associated with poorer SC-syntax (green) and SP (red) performance across all participants. Regions showing associations with both SC-syntax and SP are shown in yellow. Results are displayed at a family-wise error (FWE)-corrected threshold of p < 0.05 with a minimum cluster extent of >100 voxels. **(B)** Exploratory subtype-specific voxel-based morphometry analyses of SC-syntax performance in patients with lvPPA (blue) and nfvPPA (green). In svPPA, no clusters exceeded the predefined threshold (uncorrected voxel-wise p < 0.0001, k > 200). Abbreviations: FWE = family-wise error; lvPPA = logopenic variant primary progressive aphasia; nfvPPA = nonfluent/agrammatic variant primary progressive aphasia; PPA = primary progressive aphasia; svPPA = semantic variant primary progressive aphasia.

For SC-syntax, lower performance was associated with lower gray matter volume predominantly in a left-lateralized frontotemporal network encompassing the posterior parts of the superior, middle, and inferior temporal gyri, planum temporale, superior and middle frontal gyri, frontal operculum, inferior frontal gyrus, and anterior cingulate gyrus (green clusters, Fig. 4A; Supplementary Table 2). For SP, poorer performance was associated with lower gray matter volume primarily in the left frontal-subcortical regions, including the caudate nucleus, putamen, frontal operculum, thalamus, and hippocampus (red clusters, Fig. 4A; Supplementary Table 2). Regions showing shared associations with SC-syntax and SP included the left frontal operculum, inferior frontal gyrus, and superior temporal gyrus, as indicated by the yellow clusters in Fig. 4A.

To examine variant-specific neuroanatomical correlates of syntax processing, we performed exploratory VBM analyses separately in patients with lvPPA, nfvPPA, and svPPA for SC-syntax (Fig. 4B). These analyses were restricted to SC-syntax because the number of participants with available MRI data within each PPA variant was insufficient to support comparable analyses of SP. In lvPPA, poorer SC-syntax performance was associated with lower gray matter volume predominantly in the left superior and inferior temporal gyri, superior parietal lobule, hippocampus, and middle cingulate gyrus. In nfvPPA, poorer SC-syntax performance was associated with lower gray matter volume in the left superior and middle frontal gyri, anterior cingulate gyrus, and anterior insula. In svPPA, no clusters exceeded the predefined threshold (uncorrected voxel-wise *P* < 0.0001 with a minimum cluster extent of *k* > 200 voxels).

## Discussion

This study provides the first systematic characterization of syntax processing in Chinese-speaking individuals with PPA. Our findings demonstrate that syntax performance, as assessed by SC-syntax and SP, in Chinese reflects the combined influence of syntactic complexity and lexical composition and is also associated with semantic and repetition abilities, suggesting that syntax processing depends on lexicosemantic integration and auditory verbal short term memory in addition to grammatical computation. Consistent with this broader cognitive profile, SC-syntax and SP discriminated patients with PPA from cognitively healthy controls but provided limited discrimination between PPA variants. Neuroanatomical analyses further demonstrated that syntax processing was associated with both left-lateralized frontotemporal language networks and frontostriatal circuits, with variant-specific neuroanatomical correlates reflecting the characteristic patterns of neurodegeneration.

### Linguistic determinants of syntactic task performance

The item-level analyses demonstrated that syntactic performance in Chinese was shaped by both syntactic structure and lexical composition. Among studied linguistic factors, subject-relative clauses, passive voice constructions, and a higher proportion of function words were associated with poorer performance. Notably, subject-relative clauses were associated with poorer performance than object-relative clauses in both Chinese-speaking individuals with PPA and cognitively healthy controls. This contrasts with the canonical pattern reported in English, where object-relative clauses are generally more difficult to process than subject-relative clauses.^23–25^ One proposed explanation is that, unlike English subject-relative clauses, which preserve the canonical agent-before-theme order (e.g., “the boy who is pushing the girl“), Chinese subject-relative clauses place the modifying clause before the noun (e.g., ‘正在推女 孩的男孩’ [literally, ‘the girl-pushing boy’]), resulting in a linear sequence in which the theme (女孩, girl) precedes the agent (男孩, boy). In the absence of overt case marking, grammatical roles must be inferred primarily from word order, making such non-canonical constructions particularly demanding to process.^26–28^ Consistent with this interpretation, subject-relative clauses are also disproportionately impaired in Chinese-speaking individuals with stroke aphasia.^29–31^ The association between passive constructions and poorer sentence comprehension has likewise been observed in stroke aphasia across typologically diverse languages, including English, Dutch, German, Turkish, and Chinese.^29,32–34^ This cross-linguistic consistency suggests that passive sentences impose language-general demands on syntax processing by requiring the assignment of non-canonical thematic roles.^35^

In contrast, lexical characteristics also contributed substantially to syntax performance. Reduced function-word production in spontaneous speech is a hallmark of nfvPPA,^1^ whereas in our word-card ordering task, a higher proportion of function words was associated with poorer SP performance, likely reflecting increased demands on grammatical encoding and auditory verbal short term memory. Likewise, sentences containing more content words and greater character length were associated with poorer SC-syntax and SP performance, indicating that successful sentence processing depends not only on grammatical complexity but also on the amount of lexical information that must be maintained and integrated. Together, these findings demonstrate that syntax performance in Chinese reflects the combined influence of grammatical and lexical factors.

### Variant-specific mechanisms of syntax processing impairment in Chinese PPA

The observation that syntax performance was impaired beyond nfvPPA in our Chinese-speaking cohort has important implications for understanding the interaction between language typology and neurodegenerative disease. In predominantly English-speaking populations, grammatical impairment is considered a defining feature of nfvPPA, with sentence comprehension and production serving as key behavioural markers of the syndrome.^1,36^ By contrast, our findings suggest that syntax performance in Chinese reflects contributions from multiple cognitive systems and may therefore be impaired across several PPA variants. Our findings suggest that this difference may arise, in part, from the way syntactic complexity is encoded across languages. In morphologically rich languages, grammatical information can be encoded compactly through inflectional morphology, with distinctions such as tense, agreement, number, and case expressed through changes in word form. In contrast, Chinese, as an analytic language, expresses complex grammatical relationships primarily through additional lexical and functional elements, including function words, relative clause markers, modifiers, and other sentence constituents. Consequently, increasing syntactic complexity is typically accompanied by longer sentences containing additional lexical-semantic units that must be maintained and integrated during sentence processing, thereby increasing the cognitive demands of sentence processing beyond grammatical computation alone. Consistent with this framework, SC-syntax and SP performance were more strongly associated with semantic association and repetition measures than with motor speech performance, suggesting contributions from lexicosemantic processing and auditory verbal short term memory in addition to grammatical computation.

Our exploratory neuroanatomical findings further support this framework by demonstrating variant-specific neural correlates of syntax processing impairment. In nfvPPA, poorer SC-syntax performance was associated with lower gray matter volume in frontal regions, including the superior and middle frontal gyri, anterior cingulate cortex, and anterior insula, regions consistently implicated in grammatical processing. In contrast, poorer SC-syntax performance in lvPPA was associated predominantly with lower gray matter volume in left temporal, parietal, hippocampal, and cingulate regions, consistent with a greater contribution of auditory verbal short term memory. The involvement of posterior temporal and parietal regions, together with the observed association between SC-syntax and repetition performance, is consistent with a contribution of auditory verbal short term memory to sentence comprehension in lvPPA. Although no clusters exceeded the predefined threshold in svPPA, this negative finding may reflect the relatively small sample size for the SC-syntax analysis within this group. Together, these findings suggest that syntax processing impairments across PPA variants reflect distinct underlying cognitive and neuroanatomical mechanisms rather than a unitary grammatical deficit.^36–38^

Beyond influencing the cognitive mechanisms of sentence processing, the typological characteristics of Chinese also shape the clinical manifestation of agrammatism. As an analytic, pro-drop language, Chinese lacks obligatory inflectional markers for tense, agreement, and case while permitting omission of sentence arguments when they can be readily inferred from context. Consequently, some features commonly used to identify agrammatism in morphologically richer languages, particularly the omission or misuse of inflectional morphology, may be less informative in Chinese, while argument omission may be grammatically permissible and therefore less readily interpreted as evidence of grammatical impairment. As a result, grammatical impairment may be less overt than in morphologically rich languages and therefore more difficult to recognize. This interpretation is consistent with cross-linguistic evidence that expressive agrammatism is more prominent in Italian-speaking than English-speaking individuals with nfvPPA, likely reflecting the greater inflectional complexity of Italian.^13^ Importantly, our findings further suggest that impaired performance on sentence level tasks in Chinese should not be interpreted as evidence of grammatical impairment alone. Instead, sentence processing deficits may arise through distinct combinations of grammatical, lexical-semantic, and auditory verbal short term memory impairments depending on the PPA variant. Accordingly, assessment tools developed for Indo-European languages may not capture the manifestation of grammatical impairment in Chinese-speaking individuals with PPA. Structured sentence-level assessments may therefore be better suited to distinguish Chinese-speaking individuals with PPA from cognitively healthy controls.

### Cortical and fronto-striatal contributions to syntactic processing

Stroke aphasia and PPA provide complementary models for investigating the neuroanatomical basis of syntax processing. Whereas stroke lesions typically produce focal damage constrained by vascular territories, PPA is characterized by progressive degeneration of distributed language networks, offering a broader perspective on the neural architecture underlying sentence processing. Contemporary models derived from functional neuroimaging studies of healthy adults^39,40^ lesion-based studies of stroke aphasia,^41,42^ and functional MRI studies of PPA^8^ conceptualize syntax processing as a distributed, left-lateralized frontotemporal network rather than a single “grammar area,” with the posterior inferior frontal gyrus and posterior temporal cortex serving as its principal cortical hubs.^43–45^ Additional contributions from cingulate, parietal, and subcortical regions are thought to reflect the specific computational demands of sentence processing, including cognitive control, sequencing, and working memory.^46^ Our neuroanatomical findings are broadly consistent with this framework, demonstrating that syntax comprehension was associated predominantly with a left frontotemporal network, while syntax production additionally showed associations with frontostriatal structures. Notably, the overlap between comprehension- and production-related atrophy in the left inferior frontal gyrus further supports a central role of this region in syntax processing across modalities.

The additional involvement of the caudate nucleus, putamen, and frontal operculum during sentence production suggests an important contribution of frontostriatal circuits to sentence generation. Emerging evidence suggests that corticostriatal pathways linking Broca’s area and the caudate support syntactic sequencing and morphosyntactic processing.^47^ Furthermore, impairment in syntax processing has been reported in disorders affecting these circuits, specifically Huntington’s disease and Parkinson’s disease.^48,49^ Together, these findings support a contribution of frontostriatal circuits to syntax processing, potentially through their involvement in cognitive control and auditory verbal short term memory.

### Strengths and limitations

This study has several strengths. To our knowledge, it is the first multicentre study to systematically characterize syntax comprehension and production in a Chinese-speaking PPA cohort, extending current models of syntax processing beyond the predominantly English-speaking literature. By integrating behavioural, cognitive, and neuroanatomical analyses across multiple PPA variants, we provide complementary insights into the mechanisms underlying syntax performance and suggest that language typology may shape the relative contributions of grammatical, lexical-semantic, and auditory-verbal short-term memory processes to sentence level performance. More broadly, our findings underscore the importance of language typology in interpreting the behavioural and neural manifestations of syntax processing impairment and caution against assuming that models derived predominantly from English-speaking populations generalize directly to Chinese-speaking individuals with PPA. Recognising these language-specific mechanisms may improve the interpretation of syntax processing deficits and help avoid overattributing sentence level impairment to the nonfluent/agrammatic variant in Chinese-speaking patients.

Several limitations should be acknowledged. First, the imaging sample size within each PPA variant was modest, and the variant-specific VBM analyses should therefore be considered exploratory. Second, although the SC and SP tasks were designed to evaluate syntax processing, performance was also influenced by lexicosemantic processing and auditory verbal short term memory demands, limiting the ability to isolate grammatical mechanisms. Finally, because this study focused on Chinese-speaking individuals, the findings may not be directly generalizable to languages with different typological structures. Further cross-linguistic studies spanning diverse language families will be essential to better understand how language structure shapes the cognitive, neural, and clinical manifestations of syntax processing impairment in PPA.

## Conclusion

In Chinese-speaking individuals with PPA, syntax processing impairment extends beyond the nfvPPA and reflects contributions from grammatical computation, lexicosemantic processing, and auditory verbal short term memory. These behavioural findings are accompanied by distributed neuroanatomical associations involving left frontotemporal regions for sentence comprehension and additional frontostriatal regions for sentence production. Together, these findings suggest that the clinical and neurocognitive manifestations of syntactic impairment may depend, in part, on the structural properties of the language being processed. Language typology should therefore be considered when interpreting sentence-level deficits in PPA and when adapting diagnostic frameworks developed predominantly from English and other Indo-European languages.

## Data availability

De-identified data underlying the findings of this study are available from the corresponding author upon reasonable request and subject to institutional approval and a Material Transfer Agreement. Public data sharing is restricted because of ethical and privacy considerations related to participant confidentiality.

## Acknowledgements

We would like to thank the participants and their families for their generous contributions to this work. We are also grateful to Jana Klein, Xinge Zhao, Joanne Wu, Kevin Li, Alyssa Chen, and Yun-Ting Wang at UCSF; Clara Cheng, Tammy Lau, and Kerwin Cheung in Hong Kong; as well as Yu-Ruei Lin, Yun-Ching Lo and Yu-Chung Su in Taiwan for their invaluable assistance with data collection and entry.

## Supplementary material

Supplementary material is available at *Brain* online.

