## Supplementary Tables and Figures for "Beyond Agrammatism: Syntax Processing in Chinese Primary Progressive Aphasia"

**Supplementary Materials**

**Supplementary Table 1. Atrophy in Structures Related to Sentence Comprehension (SC)-syntax**

**Supplementary Table 2. Atrophy in Structures Related to Sentence Production (SP)**

**Supplementary Fig. 1. Examples of syntactic-foil and semantic-foil items in the sentence comprehension task**

**Supplementary Fig. 2. Univariable ROC curve analyses of SC-syntax and SP task performance.**

**Supplementary Fig. 3. Correlations between SC-syntax and SP task performance and semantic, repetition, and motor speech measures.**

**Supplementary Table 1. Atrophy in Structures Related to Sentence Comprehension (SC)-syntax**

| Regions | Voxel-level interference | | Cluster-level interference | | Peak MNI coordinates | | |
|---|---|---|---|---|---|---|---|
| | t-value | *P*-value (uncorrected) | Extent | *P*-value (FWE-corrected) | x | y | z |
| Left inferior temporal gyrus | 6.15 | <0.001 | 1180 | <0.001 | -54 | -62 | -15 |
| Left inferior temporal gyrus | 5.79 | <0.001 | | | -50 | -47 | -23 |
| Left planum temporal | 6.00 | <0.001 | 1030 | <0.001 | -48 | -35 | 6 |
| Left superior temporal gyrus | 5.89 | <0.001 | | | -57 | -38 | 3 |
| Left middle temporal gyrus | 5.51 | <0.001 | | | -53 | -59 | 15 |
| Left superior frontal gyrus medial segment | 5.98 | <0.001 | 645 | <0.001 | -14 | 32 | 30 |
| Left supplemental motor cortex | 5.85 | <0.001 | | | -12 | 15 | 41 |
| Left supplemental motor cortex | 5.52 | <0.001 | | | -11 | 6 | 45 |
| Left middle frontal gyrus | 5.94 | <0.001 | 194 | 0.001 | -30 | 38 | 38 |
| Left middle frontal gyrus | 5.31 | <0.001 | | | -38 | 36 | 30 |
| Left superior frontal gyrus | 5.86 | <0.001 | 228 | <0.001 | -21 | 53 | 20 |
| Left central operculum | 5.80 | <0.001 | 602 | <0.001 | -53 | 5 | 2 |
| Left central operculum | 5.42 | <0.001 | | | -41 | 2 | 9 |
| Left anterior cingulate gyrus | 5.71 | <0.001 | 122 | <0.001 | -14 | 45 | 3 |
| Left inferior frontal gyrus, opercular part | 5.68 | <0.001 | 494 | <0.001 | -42 | 11 | 26 |
| Left precentral gyrus | 5.60 | <0.001 | | | -47 | 3 | 29 |
| Left frontal operculum | 5.51 | <0.001 | 273 | <0.001 | -38 | 21 | 3 |
| Left middle frontal gyrus | 5.46 | <0.001 | 106 | <0.001 | -32 | 54 | -2 |
| Left medial orbital gyrus | 5.36 | <0.001 | 120 | <0.001 | -14 | 38 | -20 |

Brain regions were identified using the Neuromorphometrics (NMM) atlas. Voxel-level statistical significance was assessed voxel-wise family-wise error (FWE)-corrected significance level of P < 0.05; only clusters with an extent of >100 voxels are presented. MNI = Montreal Neurological Institute; FEW= family-wise error.

**Supplementary Table 2. Atrophy in Structures Related to Sentence Production (SP)**

| Regions | Voxel-level interference | | Cluster-level interference | | Peak MNI coordinates | | |
|---|---|---|---|---|---|---|---|
| | t-value | *P*-value (uncorrected) | Extent | *P*-value (FWE-corrected) | x | y | z |
| **Left Caudate** | 6.94 | <0.001 | 2260 | <0.001 | -4 | 12 | 0 |
| **Left frontal operculum** | 6.24 | | | <0.001 | -34 | 24 | 4 |
| **Left putamen** | 5.7 | | | <0.001 | -16 | 6 | -8 |
| **Left thalamus** | 5.89 | <0.001 | 924 | <0.001 | -22 | -33 | -3 |
| **Left hippocampus** | 5.79 | | | <0.001 | -28 | -15 | -12 |
| **Left hippocampus** | 5.53 | | | <0.001 | -20 | -14 | -12 |

Brain regions were identified using the Neuromorphometrics (NMM) atlas. Voxel-level statistical significance was assessed voxel-wise family-wise error (FWE)-corrected significance level of P < 0.05; only clusters with an extent of >100 voxels are presented. MNI = Montreal Neurological Institute; FEW= family-wise error.

**Supplementary Fig. 1. Examples of syntactic-foil and semantic-foil items in the sentence comprehension task**

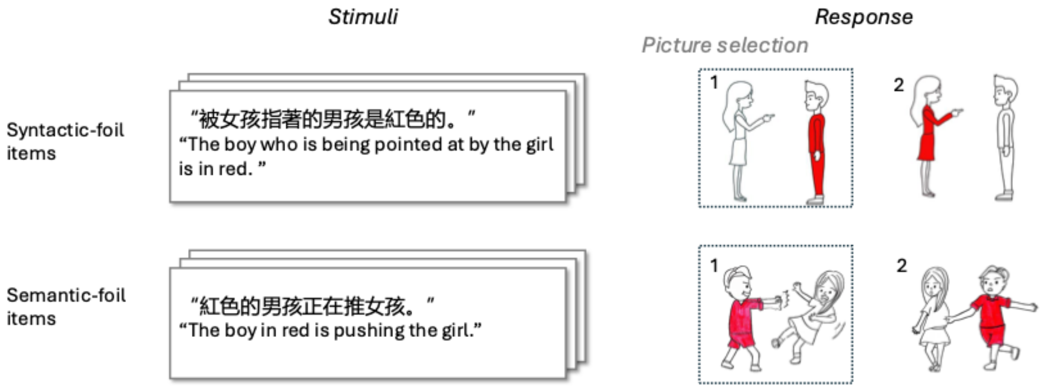


For syntactic-foil items, the picture alternatives differ in how the sentence structure relates the characters and their attributes, requiring comprehension of word order and syntactic relationships. For semantic-foil items, the picture alternatives differ in the semantic information conveyed by the sentence. In each example, participants selected the picture that best matched the sentence.

**Supplementary Fig. 2. Univariable ROC curve analyses of SC-syntax and SP task performance.**

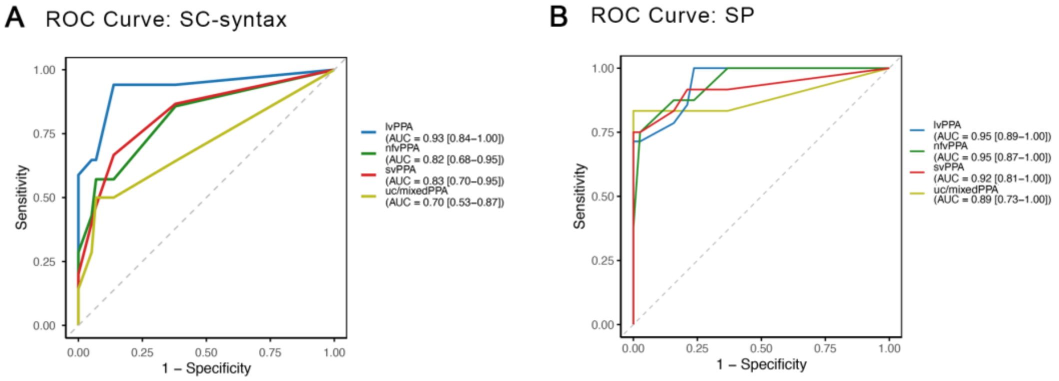


Unadjusted ROC curves showing the discriminative performance of SC-syntax (A) and SP (B) tasks in distinguishing each PPA variants from cognitively normal controls.

ROC = Receiver Operating Characteristic, SC = sentence comprehension, SP = sentence production

**Supplementary Fig. 3. Correlations between SC-syntax and SP task performance and semantic, repetition, and motor speech measures.**

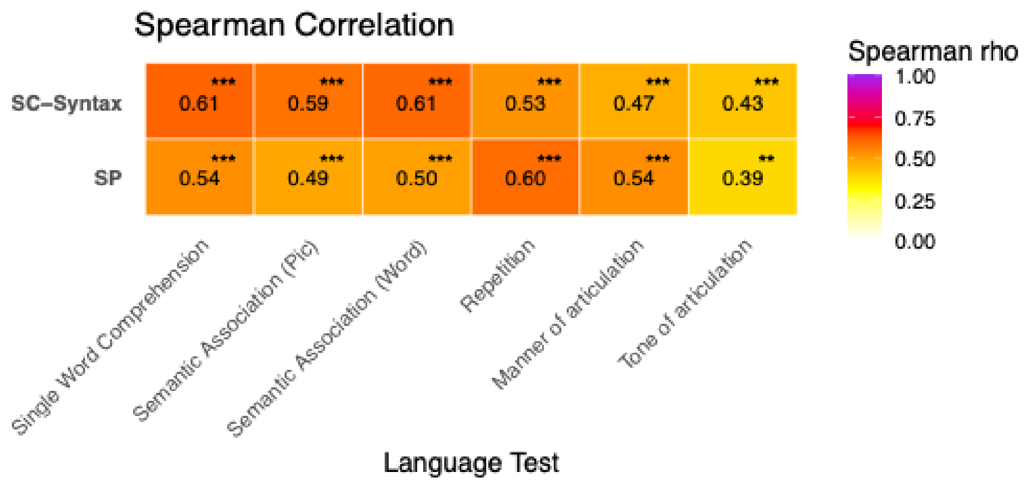


Values represent Spearman rank correlation coefficients (ρ) between SC-syntax, SP, and language/cognitive test scores. ** $P < 0.01$; *** $P < 0.001$.

SC = sentence comprehension, SP = sentence production
